# Validation of the BOADICEA Model in a Prospective Cohort of *BRCA1/2* Pathogenic Variant Carriers

**DOI:** 10.1101/2024.04.21.24306136

**Authors:** Xin Yang, Thea M. Mooij, Goska Leslie, Lorenzo Ficorella, Nadine Andrieu, Karin Kast, Christian F. Singer, Anna Jakubowska, Carla H. van Gils, Yen Tan, Christoph Engel, Muriel A. Adank, Christi J. van Asperen, Margreet G.E.M. Ausems, Pascaline Berthet, EMBRACE collaborators, J. Margriet Collée, Jackie Cook, Jacqueline Eason, K.Y. van Spaendonck-Zwarts, D Gareth Evans, Encarna B. Gomez Garcia, Helen Hanson, Louise Izatt, Zoe Kemp, Fiona Lalloo, Christine Lasset, Fabienne Lesueur, Hannah Musgrave, Sophie Nambot, Catherine Noguès, Jan C. Oosterwijk, Dominique Stoppa-Lyonnet, Marc Tischkowitz, Vishakha Tripathi, Marijke R. Wevers, Emily Zhao, Flora van Leeuwen, Marjanka K. Schmidt, Douglas F. Easton, Matti A. Rookus, Antonis C. Antoniou

## Abstract

**Background:** No validation has been conducted for the BOADICEA multifactorial breast cancer risk prediction model specifically in *BRCA1/2* pathogenic variant (PV) carriers to date. Here, we evaluated the performance of BOADICEA in predicting five-year breast cancer risks in a prospective cohort of *BRCA1/2* PV carriers ascertained through clinical genetic centres.

**Methods:** We evaluated the model calibration and discriminatory ability in the prospective TRANsIBCCS cohort study comprising 1,614 *BRCA1* and 1,365 *BRCA2* PV carriers (209 incident cases). Study participants had lifestyle, reproductive, hormonal, anthropometric risk factor information, a polygenic risk score based on 313 single nucleotide polymorphisms, and family history information.

**Results:** The full multifactorial model considering family history together with all other risk factors was well calibrated overall (E/O=1.07, 95%CI:0.92-1.24) and in quintiles of predicted risk. Discrimination was maximized when all risk factors were considered (Harrell’s C-index=0.70, 95%CI:0.67-0.74; AUC=0.79, 95%CI:0.76-0.82). The model performance was similar when evaluated separately in *BRCA1* or *BRCA2* PV carriers. The full model identified 5.8%, 12.9% and 24.0% of *BRCA1/2* PV carriers with five-year breast cancer risks of <1.65%, <3% and <5% respectively, risk thresholds commonly used for different management and risk-reduction options.

**Conclusion:** BOADICEA may be used to aid personalised cancer risk management and decision making for *BRCA1* and *BRCA2* PV carriers. It is implemented in the free-access CanRisk tool (www.canrisk.org).

**What is already known on this topic:** No study has assessed the clinical validity of the multifactorial BOADICEA model for predicting future breast cancer risks specifically for *BRCA1/2* pathogenic variant (PV) carriers.

**What this study adds:** This is the first study to validate the BOADICEA model based on the joint effects of questionnaire-based risk factors (QRFs), a polygenic risk score (PRS) based on 313 single nucleotide polymorphisms, and cancer family history information in *BRCA1/2* PV carriers ascertained through clinical genetic centres. The model is well calibrated and discriminated well in both *BRCA1* and *BRCA2* carriers. The inclusion of family history, alongside QRFs and the PRS, in predicting cancer risks for PV carriers in clinical genetics settings can improve the calibration within individual risk categories and can result in clinically meaningful levels of breast cancer risk stratification.

**How this study might affect research, practice or policy:** BOADICEA is freely available via the CanRisk tool (www.canrisk.org). Rather than relying solely on average published penetrance estimates commonly used in genetic clinics for counselling of *BRCA1/2* PV carriers, BOADICEA offers more personalized BC risks. This can facilitate informed decision-making regarding the clinical management of BC risk, including considerations for surveillance and the timing of risk-reducing surgery.

## Introduction

Women with pathogenic variants (PVs) in *BRCA1* and *BRCA2* (henceforth called “PV carriers”) are at high risk of developing breast (BC) and ovarian cancer[1]. However, BC risks for PV carriers vary by family history (FH) and by other genetic, lifestyle, hormonal and reproductive factors which can result in variability in the individualised BC risk assessment[2-5]. Providing more personalized BC risks will enable informed decision-making for the clinical management of BC risk, for example opting for bilateral risk-reducing mastectomy and its timing.

The BOADICEA model, implemented in the CanRisk tool (www.canrisk.org), predicts the risk of developing BC by considering the combined effects of rare genetic variants in *BRCA1, BRCA2, PALB2, CHEK2, ATM, RAD51C, RAD51D*, and *BARD1*, a polygenic risk score (PRS), FH, mammographic density (MD), and questionnaire-based risk factors (QRFs) including hormonal, lifestyle and reproductive factors[6, 7]. Previous validation studies in independent prospective cohorts have shown that the model is well calibrated and provides good discrimination in the general population[8-10]. However, the model performance has not been evaluated specifically in *BRCA1/2* PV carriers. Here, we evaluate the performance of BOADICEA v.6[7] in predicting BC risks in an independent prospective cohort of *BRCA1* and *BRCA2* PV carriers.

## Methods

### Subjects

Data on 2,879 *BRCA1* and 2,208 *BRCA2* female PV carriers were available from the prospective TRANsIBCCS cohort study[11]. Participants were recruited via clinical genetics centres in Germany (GC-HBOC), the UK (EMBRACE), France (GENEPSO), the Netherlands (HEBON), Austria (MUV) and Poland (IHCC) and were counselled with regard to their mutation status. All participants were heterozygotes of variants considered to be pathogenic on the basis of widely accepted criteria (ENIGMA consortium; https://enigmaconsortium.org/).

All the participants were actively followed up for cancer incidence and mortality through follow-up questionnaires. In addition, follow-up through linkage with cancer, pathology and death registries has been provided in countries where these registries are available (cancer/death registries in the Netherlands and the UK; pathology registries to collect information on preventive surgeries in the Netherlands, and through medical record validation of self-reported preventive surgeries) [11]. Study protocols were approved by local ethics committees and all individuals gave informed consent.

### Censoring process

All participants were followed from age at baseline to the date of BC diagnosis (invasive or ductal carcinoma in situ (DCIS)), bilateral risk-reducing mastectomy, last follow-up, death, baseline plus six years or age 80, whichever occurred first. Only those with a BC diagnosis were considered affected. A total of 344 women were censored at bilateral prophylactic mastectomy.

### Risk prediction, model calibration and discrimination

To exclude potentially prevalent but undiagnosed BC patients at study recruitment, we predicted the five-year BC risks starting from the age at study entry plus one year. The study used the latest version of BOADICEA v.6 [7] implemented in CanRisk version 2.4 (https://canrisk.org/releases/) [12]. We evaluated the model calibration and discriminatory ability. The overall calibration was assessed by the ratio of the expected (E) to the observed (O) number of incident BC patients during the five-year risk prediction period [13]. We also assessed the agreement between predicted and observed risk for each individual using the calibration slope, which was calculated by fitting a logistic regression in which the dependent variable was the observed outcome (1:affected; 0:unaffected) and the independent variable was the log odds of the predicted risks. The calibration slope assesses whether the predicted risks are too extreme or conversely too moderate especially at the high and low risk tails and is expected to be equal to 1 if the model is perfectly calibrated. The observed and expected risks were also compared in categories by grouping the samples in quintiles of predicted risks. Discrimination was assessed by the area under the receiver operating characteristic curve (AUC) and Harrell’s C-index[14]. To assess the risk-stratifying ability of the model, we calculated the proportions of all women that had five-year BC risks of <1.65%, <3% or <5% which are the commonly used thresholds for discussing risk-reducing options[15, 16] and also examined the proportion of women younger than 50 years old in the low-risk groups who may opt out of or delay the risk-reducing surgeries.

From the total of 5,087 women in the entire TRANsIBCCS prospective cohort, women were selected for inclusion in the analysis if they were younger than 74 years old at study-entry, if they had no history of cancer or bilateral risk-reducing mastectomy, had more than one year follow-up and had data on QRFs and the 313-SNP PRS [17] (Figure s1). The 313-PRS was standardised using a mean of - 0.424 and standard deviation of 0.611 as described in [17]. Models were then evaluated in: (1) the cohort of 2,979 women who had QRF and PRS data (cohort-1); (2) among those, a cohort of 1,804 women with QRF, PRS and pedigree-based cancer family history information available (cohort-2). To allow for the possibility that inclusion in these two sub-cohorts is non-random with respect to the incident BC status compared to the entire TRANsIBCCS prospective cohort, sampling weights were applied to the final set of eligible women in each sub-cohort. The sample inclusion probabilities were computed by fitting a logistic regression model in which the outcome (inclusion or not) was dependent on the age at baseline, follow-up duration, incident BC status, the interaction between BC status, age at baseline and the interaction between BC status and the follow-up duration. These were calculated for each country separately, except for Austria, Germany and Poland which were combined due to the limited sample size. The weights were then the inverse of the fitted probabilities for each individual.

All the statistical analyses were performed in R version 3.6.3[18].

## Results

A total of 2,979 European ancestry *BRCA1/2* PV carriers with information on PRS and QRFs were eligible for inclusion in the analysis, of whom 209 (127 *BRCA1* and 82 *BRCA2* PV carriers) developed BC during the five-year risk prediction period (cohort-1). Among these, 1,804 women (191 with incident BC) also had pedigree-based FH (cohort-2). A detailed summary of the genetic and epidemiological characteristics of the study participants at baseline are shown in Table s1. We evaluated the model separately in cohort-1 without considering FH and in cohort-2 considering the pedigree-based FH information.

Using cohort-1, when considering *BRCA1* and *BRCA2* PV status only, or *BRCA1* and *BRCA2* PV status and QRFs, the predicted risks were underestimated (Table 1) in particular for women in the higher predicted risk quintiles (Figure 1A). The addition of PRS to PV status improved the calibration of the predicted risks (E/O=0.88, 95%CI:0.76-1.01, calibration slope=0.95, 95%CI:0.90-1.00, Figure 1A). Similarly, adding PRS to the model with PV and QRFs information improved calibration, but discrimination was similar (Table 1).

**Table 1:** Calibration and discrimination of five-year predicted breast cancer risks under the BOADICEA model using different risk factor combinations.

| Model | Category | AUC | Harrell's C-index | E/O | Calibration slope |
| --- | --- | --- | --- | --- | --- |
| **using cohort-1, N=2979 including 209 incident BCs (*BRCA1*: 1614 including 127 incident BCs; *BRCA2*: 1365 including 82 incident BCs)** |  |  |  |  |  |
| Null (age only) | All women | 0.70 (0.66, 0.73) | 0.64 (0.59, 0.67) | 0.06 (0.05, 0.07) | 0.45 (0.42, 0.47) |
|  | *BRCA1* PV carriers | 0.69 (0.64, 0.74) | 0.62 (0.57, 0.67) | 0.05 (0.04, 0.06) | 0.42 (0.39, 0.44) |
|  | *BRCA2* PV carriers | 0.72 (0.67, 0.78) | 0.67 (0.61, 0.74) | 0.08 (0.06, 0.10) | 0.49 (0.45, 0.53) |
| PV | All women | 0.76 (0.73, 0.80) | 0.68 (0.64, 0.72) | 0.80 (0.69, 0.93) | 0.93 (0.88, 0.98) |
|  | *BRCA1* PV carriers | 0.75 (0.71, 0.79) | 0.65 (0.59, 0.70) | 0.83 (0.69, 1.00) | 0.94 (0.87, 1.01) |
|  | *BRCA2* PV carriers | 0.79 (0.74, 0.84) | 0.73 (0.67, 0.77) | 0.75 (0.60, 0.95) | 0.92 (0.85, 1.00) |
| PV+QRFs | All women | 0.78 (0.76, 0.81) | 0.69 (0.66, 0.74) | 0.78 (0.67, 0.90) | 0.93 (0.88, 0.98) |
|  | *BRCA1* PV carriers | 0.77 (0.73, 0.81) | 0.67 (0.61, 0.72) | 0.82 (0.68, 0.98) | 0.94 (0.87, 1.01) |
|  | *BRCA2* PV carriers | 0.81 (0.76, 0.85) | 0.74 (0.68, 0.79) | 0.72 (0.57, 0.91) | 0.91 (0.84, 0.99) |
| PV+PRS | All women | 0.77 (0.73, 0.80) | 0.68 (0.64, 0.71) | 0.88 (0.76, 1.01) | 0.95 (0.90, 1.00) |
|  | *BRCA1* PV carriers | 0.75 (0.70, 0.79) | 0.66 (0.59, 0.70) | 0.91 (0.75, 1.09) | 0.95 (0.88, 1.02) |
|  | *BRCA2* PV carriers | 0.79 (0.74, 0.83) | 0.72 (0.68, 0.78) | 0.83 (0.66, 1.05) | 0.95 (0.87, 1.03) |
| PV+QRFs+PRS | All women | 0.78 (0.75, 0.81) | 0.69 (0.66, 0.73) | 0.86 (0.74, 0.99) | 0.95 (0.89, 1.00) |
|  | *BRCA1* PV carriers | 0.76 (0.72, 0.80) | 0.66 (0.62, 0.72) | 0.89 (0.74, 1.07) | 0.95 (0.88, 1.02) |
|  | *BRCA2* PV carriers | 0.80 (0.76, 0.84) | 0.73 (0.69, 0.78) | 0.80 (0.64, 1.01) | 0.94 (0.86, 1.01) |
| **using cohort-2, N=1804 including 191 incident BCs (*BRCA1*: 1016 including 118 incident BCs; *BRCA2*: 788 including 73 incident BCs)** |  |  |  |  |  |
| PV+QRFs+PRS | All women | 0.78 (0.75, 0.81) | 0.69 (0.65, 0.72) | 0.85 (0.74, 0.99) | 0.94 (0.89, 1.00) |
|  | *BRCA1* PV carriers | 0.76 (0.72, 0.80) | 0.67 (0.62, 0.72) | 0.87 (0.72, 1.05) | 0.95 (0.88, 1.02) |
|  | *BRCA2* PV carriers | 0.78 (0.74, 0.83) | 0.72 (0.67, 0.78) | 0.83 (0.65, 1.06) | 0.94 (0.86, 1.02) |
| FH+QRFs+PRS+PV | All women | 0.79 (0.76, 0.82) | 0.70 (0.67, 0.74) | 1.07 (0.92, 1.24) | 1.06 (1.00, 1.12) |
|  | *BRCA1* PV carriers | 0.78 (0.74, 0.82) | 0.69 (0.62, 0.74) | 1.05 (0.87, 1.27) | 1.05 (0.97, 1.13) |
|  | *BRCA2* PV carriers | 0.79 (0.75, 0.84) | 0.72 (0.66, 0.77) | 1.10 (0.86, 1.40) | 1.07 (0.98, 1.16) |

PV: pathogenic variant status in *BRCA1* and *BRCA2*; QRFs: questionnaire-based risk factors; PRS: polygenic risk score; FH: family history.

**Figure 1:**
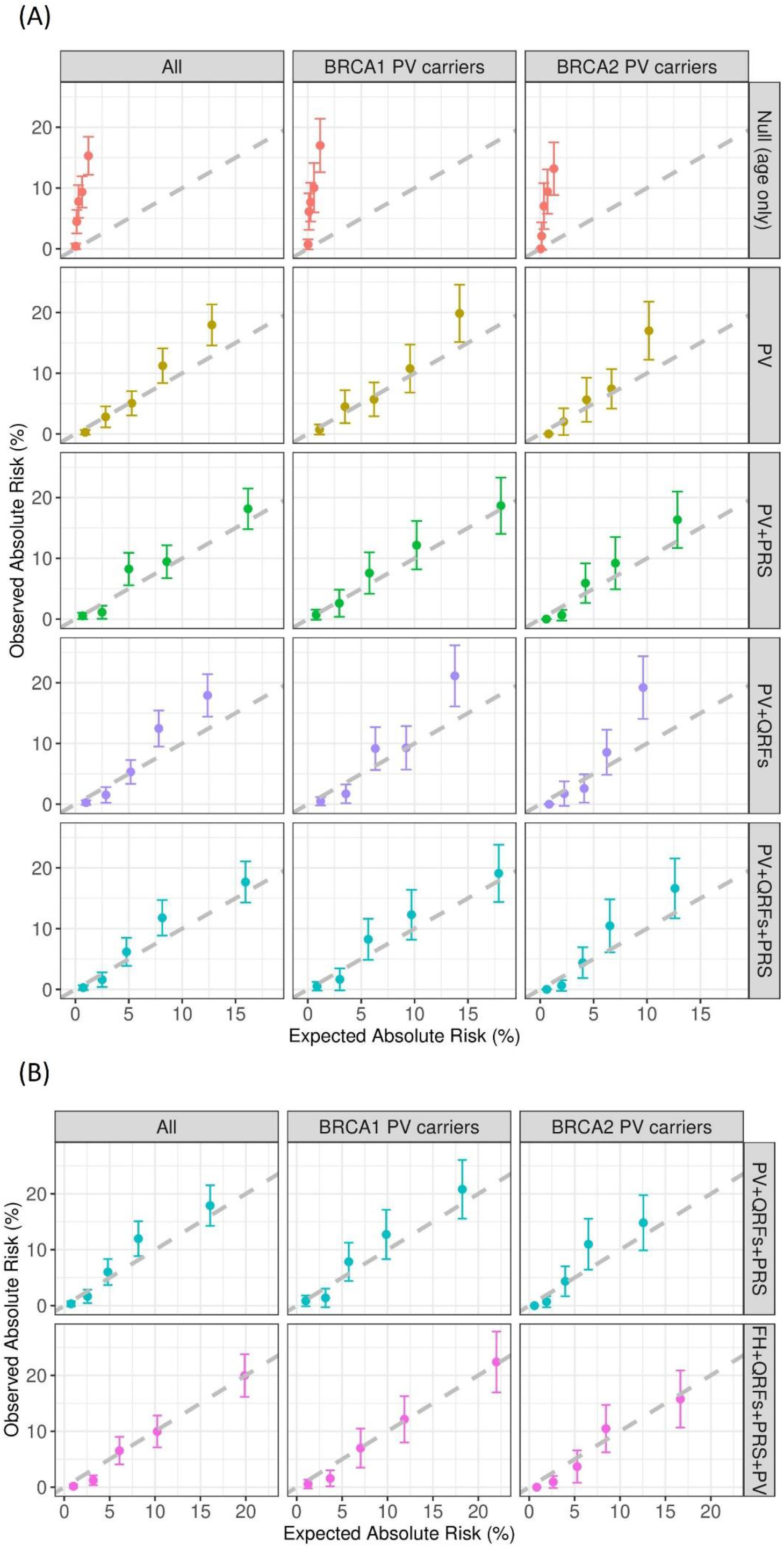
Observed and expected five-year breast cancer risks in quintiles of predicted risks.(A) using the cohort-1 samples (N=2,979) under the models considering Null (age only), PV, PV+PRS, PV+QRFs and PV+QRFs+PRS; (B) using the cohort-2 samples with FH information (N=1,804) under the models considering PV+QRFs+PRS and FH+QRFs+PRS+PV. The dashed line is the diagonal line with slope equal to 1 (corresponding to E/O ratio of 1 for each quintile). PV: pathogenic variant status in *BRCA1* and *BRCA2*; QRFs: questionnaire-based risk factors; PRS: polygenic risk score; FH: family history.

Using cohort-2, we first assessed the model predictions by leaving FH out to contrast against the results in cohort-1. The model discriminatory ability and model calibration were similar to the estimates using all 2,979 samples (Table 1). These suggest that no bias was introduced when using the weighting cohort approach in analysing the data. After including full pedigree-FH information in the model five-year risk predictions, the model was well calibrated (overall E/O=1.07, 95%CI:0.92-1.24; calibration slope=1.06, 95%CI:1.00-1.12, Table 1, Figure 1B). There was a small increase in the model discriminatory ability (Harrell’s C=0.70, 95%CI:0.67-0.74; AUC=0.79, 95%CI:0.76-0.82, Table 1). The model performance was similar in *BRCA1* and *BRCA2* PV carriers (Table1, Figure 1B).

When considering all risk factors jointly, the predicted five-year risks varied from 0.1% to 47.6%. A total of 5.8%, 12.9% and 24.0% of women had five-year BC risks of <1.65%, <3% and <5% with a negative predictive value at the 5% risk-threshold of 0.96 (95%CI:0.96-0.97). 98.0% of women with a five-year BC risk of 3% or lower, and 95.7% women (including all *BRAC1* PV carriers and 91.5% of *BRCA2* PV carriers) with a five-year BC risk of 5% or lower were younger than 50 years old. Among women younger than 50 years old, 98.5% with five-year risk of <5%, remained unaffected during the risk prediction period. Furthermore, 78.4% of women younger than 30 years old were predicted to have five-year risk of <5%; among them, 99.2% remained unaffected during the risk prediction period.

## Discussion

Previous validation studies have demonstrated that BOADICEA provides valid BC risks for women in the general population or women participating in screening programmes[8-10]. Since *BRCA1/2* PVs are rare in the population, it has not been possible to assess the model performance specifically in PV carriers who are typically seen in clinical genetics[10]. Although previous studies have indicated that multiple risk factors modify the BC risks for PV carriers[2, 19-22], their combined effects on risk prediction[1] has not been studied. Here, for the first time, we examined the model performance of the multifactorial BOADICEA model in predicting BC risks in *BRCA1* and *BRCA2* PV carriers seen at clinical genetics using information on PV, PRS, QRFs and FH jointly and showed that the BOADICEA is well calibrated and discriminated in this population. The results suggest that considering FH when predicting cancer risks for PV carriers seen in clinical genetics, in addition to QRFs and the PRS, can improve the calibration within individual risk categories. Given the majority of such women come from families with cancer FH, and the FH distribution in this cohort is not representative of the distribution in the general population, ignoring FH can result in some underprediction of risk among those who are at higher risk. Therefore, considering only average, published penetrance estimates for the counselling of *BRCA1/2* PV carriers typically seen in genetic clinics may underestimate BC risks – a scenario equivalent to the predictions in cohort-1, when using only *BRCA1* and *BRCA2* PV status. The analyses considered the full pedigree-based FH collected, which included third degree or more distant relatives. When the analysis was restricted to include only first or second degree relatives the model performance was comparable (Table s2 and Figure s2), indicating the collection of less extensive family history may be cost-effective in clinical risk assessment.

Here, in the cohort of *BRCA1* and *BRCA2* PV carriers, the AUC of 0.79 (95%CI=0.76-0.82) is higher than estimates from validation studies in population-based cohorts[8-10]. Terry et al. using multigenerational pedigree data from Australia, Canada, and the USA [23] showed that a previous version of BOADICEA that considered FH and PV status only had a C-index of 0.59 and overpredicted the 10-year risk for combined *BRCA1* and *BRCA2* PV carriers in the highest quintile. However, the study used an older version of BOADICEA (v.3). Here, we used the latest model [7, 12], and the analysis included additional risk factors (e.g. QRFs and PRS). These, together with the differences in the risk prediction period, the age distributions and other cohort characteristics, makes a direct comparison difficult. The present study suggests that the latest model is well calibrated across different risk categories in *BRCA1* and *BRCA2* carriers.The AUC estimates here could potentially have been overestimated because the risks for healthy women were predicted to the censoring age if they were censored within the risk prediction period. To address this we also estimated and presented the Harrell’s C-index[14] which considers time-to-event. The Harrell’s C-index yielded lower estimates than the AUC for all models. The full model that jointly considered all risk factors provided the highest discrimination as measured by Harrell’s C-index (Table 1). Another potential explanation of the higher discriminatory ability observed in the current study is most likely due to the larger effect of age on BC risks for *BRCA1* and *BRCA2* PV carriers compared with the general population and the age-range of study participants in this study. When only age was considered in the model, the estimated AUC in the present cohort of *BRCA1* and *BRCA2* PV carriers was 0.70 (95%CI:0.66-0.73) much higher than the effect of age alone in population-based studies[10] (Figure 1A, Table 1, cohort-1).

The changes in the C-index (or AUC) by the inclusion of additional risk factors on top of PV status are not significant, based on the associated confidence intervals. This could be a consequence of the relatively small sample size. Nevertheless, the full model, that includes PV status, FH, PRS and QRFs has the highest C-index. Given the high BC risks for *BRCA1/2* PV, even modest increases in the C-index can lead to changes in risk stratification[10]. For example, when considering the half of the PV carriers with the highest predicted risks, the full model identifies 91.2% of incident BCs occurring during the prediction period. This compares to identifying 82.2% of incident BCs when only age and pathogenic variant are considered. Moreover, the observed variability in the BOADICEA predicted risks suggests that it is possible to identify *BRCA1* and *BRCA2* PV carriers with relatively low risks, in particular among women under 50 years old (or women under 30), who remain disease free during the five year period. The results suggest that during the genetic counselling process, considering the joint effects of risk factors could be informative for decisions on the timing of risk-reducing interventions

Analysis was repeated by censoring women diagnosed with DCIS as unaffected at the age at diagnosis (Table s3, Figure s3). The model discriminatory ability as measured by the AUC remained similar to the overall analyses, when DCIS were considered as affected, as expected there was some increase in the ratio of E/0 cases (1.18; 95%CI:1.01 -1.38) and the calibration slope (1.11, 95%CI:1.05-1.17) for the full model using cohort-2, suggesting some overall overprediction of risks. However, the model was still well calibrated within quintiles of predicted risk, with no significant differences between the observed and predicted risk (Figure s3).

BOADICEA does not consider the potential effect of risk-reducing salpingo-oophorectomy (RRSO) on BC risk. Censoring at RRSO resulted in some miscalibration in quintiles of predicted risk (Figure s4). Previous studies have shown that mammographic density (MD) is also a risk factor for BC in *BRCA1* and *BRCA2* PV carriers[24, 25]. Although BOADICEA considers the effect of MD in predicting BC risks, the number of women with MD data at baseline was too small (N=794) to allow for a model assessment, which is a major limitation of the study. The number of *BRCA1* and *BRCA2* carriers was relatively small when divided by age. Nevertheless, when assessed separately by age 50, the full model was well calibrated in the <50 years age group. There was some overprediction in women aged 50 years or older with E/O ratio of 1.28 (95%CI:0.96, 1.71) but this was not significant (Figure s5, Table s4). Larger number of carriers at older ages, with a larger number of incident cancers will be required to assess the predicted risks with greater precision, in particular among different risk categories [26]. BOADICEA assumes that the joint effects of *BRCA1/2* PVs with the PRS and QRFs are multiplicative on the risk scale [6, 7] but studies suggested that deviations from the multiplicative model may exist.[2, 5] BOADICEA models assumes an age-dependent effect of the PRS, as previously described[2] and the present study suggests the BOADICEA assumptions, provide valid risks for *BRCA1/2* PV carriers. Much larger sample sizes will be required to detect small deviations between the observed and predicted risk.

In conclusion, in the overall TRANsIBCCS prospective cohort of *BRCA1* and *BRCA2* PV carriers who were ascertained through genetic clinics, primarily on the basis of cancer FH, the multifactorial BOADICEA provided good discriminatory ability and was calibrated in predicting five-year risks within different risk categories. The results suggest that BOADICEA may be used to aid personalised cancer risk management and decision making for *BRCA1* and *BRCA2* PV carriers. However, the number of PV carriers by country was too small to assess differences in the predictive ability of the model by country or to assess how potential differences in data collection practices for outcomes and elective surgeries by country/study may influence the results. Future studies with much larger sample sizes of *BRCA1* and *BRCA2* PV carriers by country and with long-term follow-up should be performed to assess BOADICEA. Furthermore, it will be important to assess whether the prediction performance can be improved by using *BRCA1/2* specific parameter estimates for the effects of the PRS and QRFs in the model.

## Data Availability

Data may be made available upon request and after review of the study proposal by the International BRCA1/2 Carrier Cohort Study (IBCCS) Data Access Coordinating Committee; please contact for further information.

## Data Availability Statement

Data may be made available upon request and after review of the study proposal by the International *BRCA1/2* Carrier Cohort Study (IBCCS) Data Access Coordinating Committee; please contact for further information.

## Acknowledgements

This work was supported by TRANsIBCCS JT (2021/cancer12-054); NKI2013-6403 and by grants from Cancer Research UK (C12292/A20861 and PPRPGM-Nov20\100002) and by the NIHR Cambridge BRC. The national French cohort, GENEPSO, was initially funded by grants from the Fondation de France and the Ligue Nationale Contre le Cancer and is being supported by Institut National du Cancer-DGOS, grant PRT-K22-076 and by the “Programmes labellisés (PGA) 2022” of the Fondation ARC (N°ARCPGA2022010004414_4863). It also received support from the European commission FP6 (Project GEN-RAD-RISK 2005) and the French National Institute of Cancer (INCa SHS-E-SP 2008, 2011, 2015; CANSOP 2014). GENEPSO is supported for this project by an INCa grant as part of the European program ERA-NET on Translational Cancer Research (TRANsIBCCS-JTC2012, no. 2014-008). The GEMO biobank was initially funded by the French National Institute of Cancer (INCa, PHRC Ile de France, grant AOR 01 082, 2001-2003, grant 2013-1-BCB-01-ICH-1), the Association “Le cancer du sein, parlons-en” Award (2004), the Association for International Cancer Research (2008-2010), and the Fondation ARC pour la recherche sur le cancer (grant PJA 20151203365). It also received support from the Canadian Institute of Health Research for the “CIHR Team in Familial Risks of Breast Cancer” program (2008-2013), and the European commission FP7, Project «Collaborative Ovarian, breast and prostate Gene-environment Study (COGS), Large-scale integrating project» (2009-2013). GEMO is currently supported by the INCa grant SHS-E-SP 18-015. MT was supported by the NIHR Cambridge Biomedical Research Centre (BRC-1215-20014). The International Hereditary Cancer Centre (IHCC) was supported by Grant PBZ_KBN_122/P05/2004 and The National Centre for Research and Development (NCBR) within the framework of the international ERA-NET TRANSAN JTC 2012 application no. Cancer 12-054 (Contract No. ERA-NET-TRANSCAN/07/2014). The HEBON study is supported by the Dutch Cancer Society grants NKI1998-1854, NKI2004-3088, NKI2007-3756, NKI 12535, the Netherlands Organisation of Scientific Research grant NWO 91109024, the Pink Ribbon grants 110005 and 2014-187.WO76, the BBMRI grant NWO 184.021.007/CP46, and the Transcan grant JTC 2012 Cancer 12-054.MT was supported by the NIHR Cambridge Biomedical Research Centre (BRC-1215-20014).

We acknowledge the GENEPSO Coordinating Center: DRCI, Institut Paoli-Calmettes, Marseille, France: Catherine Noguès, MD, Lilian Laborde, Emanuelle Breysse, Anne Robert-Bourgoin, Ulysse Bousquet, Pauline Heux; the genetic epidemiology platform (the PIGE, Plateforme d’Investigation en Génétique et Epidémiologie, Institut Curie, Paris) and particularly Juana Beauvallet who centralised, digitalized mammograms and coded and computed pedigrees data for the TRANsIBCCS Project. We acknowledge the GENEPSO Collaborating Centres and Investigators. We also acknowledge investigators of the Genetic Modifiers of Cancer Risk in BRCA1/2 Mutation Carrier (GEMO) study from the National Cancer Genetics Network UNICANCER Genetic Group, France. GEMO contributed in providing genotype data of the GENEPSO participants thanks to its participation to CIMBA consortium. We thank Noura Mebirouk who managed the GEMO samples, Sandrine Caputo who maintains the French *BRCA1*/*2* variants database and helped verifying variants nomenclature and classification, and Yue Jiao who developed the record linkage process to make GEMO and GENEPSO databases interoperable.

The Hereditary Breast and Ovarian Cancer Research Group Netherlands (HEBON) consists of the following Collaborating Centers: Netherlands Cancer Institute (coordinating center), Amsterdam, Netherlands (NL); Erasmus Medical Center, Rotterdam, NL; Leiden University Medical Center, NL; Radboud University Medical Center Nijmegen, NL; University Medical Center Utrecht, NL; Amsterdam UMC, University of Amsterdam, NL; Amsterdam UMC, Vrije Universiteit Amsterdam, NL; Maastricht University Medical Center, NL; University of Groningen, NL; The Netherlands Comprehensive Cancer Organisation (IKNL; The nationwide network and registry of histo- and cytopathology in The Netherlands (PALGA). HEBON thanks all study participants. GC-HBOC is supported for this project by a BMBF grant (01KT1405) as part of the European program ERA-NET on Translational Cancer Research (TRANsIBCCS-JTC2012, no. 2014-008). The GC-HBOC is supported by the German Cancer Aid (grant no 110837 and grant no 70114178, coordinator: Rita K. Schmutzler, Cologne) and the Federal Ministry of Education and Research (BMBF), Germany (grant no 01GY1901).

## Author Contributions

Conceptualization: M.A.R., A.C.A., D.F.E.; Data curation: T.M.M., G.L.; Formal analysis: X.Y.; Supervision: M.A.R., A.C.A.; Visualization: X.Y.; Writing-original draft: X.Y., A.C.A., M.A.R., D.F.E.; Writing-review & editing: all authors.

## Ethics Declaration

This study was approved by local ethics committees and all individuals gave informed consent. TRANsIBCCS consists six study centres including GC-HBOC, EMBRACE, GENEPSO, HEBON, MUV and IHCC. GC-HBOC was approved by the Ethics Commission of Cologne University’s Faculty of Medicine (07/048), the Ethics Commission of Dresden University’s Faculty of Medicine (EK 205052015), the Ethics Commission of the Technical University Munich, Rechts der Isar, Faculty of Medicine (169/15s), the Ethics Commission of Düsseldorf University’s Faculty of Medicine (4884), the Ethics Commission of Kiel University’s Faculty of Medicine, the Ethic Commission of Hannover Medical School (Nr. 4121), the Ethics Commission of Münster University’s Faculty of Medicine. EMBRACE was approved by the East of England - Cambridge South Research Ethics Committee (MREC 98/5/27). GENEPSO was approved by the Commission Nationale de l’Informatique et des Libertés (CNIL agreement N°999350V4-2017). The genetic data for GENEPSO participants are collected via the partner GEMO study. GEMO was reviewed by the Comité consultatif sur le traitement de I’information en matière de recherche dans le domaine de la santé (CCTIRS N°07223) and Commission Nationale de l’Informatique et des Libertés (CNIL N°1245228) and was considered compliant with the General Data Protection Regulation and was approved by the institutional review committee of Institut Curie on April 2020. HEBON was approved by the Protocol Toetsingscommissie van het Nederlands Kanker Instituut/Antoni van Leeuwenhoek Ziekenhuis (PTC07.1611/2007NKI-2007-375). MUV was approved by the Ethikkommission der Medizinischen Universität Wien (2190/2019). IHCC was approved by the Komisja Bioetyczna Pomorskiego Uniwersytetu Medycznego w Szczecinie (Bioethics Committee of Pomeranian Medical University in Szczecin; BN-001/33/04).

## Conflict of Interest

ACA and DFE are named creators of the BOADICEA model which has been licensed by Cambridge Enterprise (University of Cambridge). All the other authors declare no conflict of interest.

## Notes

### Author Declarations

This study was approved by local ethics committees and all individuals gave informed consent. TRANsIBCCS consists six study centres including GC-HBOC, EMBRACE, GENEPSO, HEBON, MUV and IHCC. GC-HBOC was approved by the Ethics Commission of Cologne University’s Faculty of Medicine (07/048), the Ethics Commission of Dresden University’s Faculty of Medicine (EK 205052015), the Ethics Commission of the Technical University Munich, Rechts der Isar, Faculty of Medicine (169/15s), the Ethics Commission of Düsseldorf University’s Faculty of Medicine (4884), the Ethics Commission of Kiel University’s Faculty of Medicine, the Ethic Commission of Hannover Medical School (Nr. 4121), the Ethics Commission of Münster University’s Faculty of Medicine. EMBRACE was approved by the East of England - Cambridge South Research Ethics Committee (MREC 98/5/27). GENEPSO was approved by the Commission Nationale de l’Informatique et des Libertés (CNIL agreement N999350V4-2017). The genetic data for GENEPSO participants are collected via the partner GEMO study. GEMO was reviewed by the Comité consultatif sur le traitement de I’information en matière de recherche dans le domaine de la santé (CCTIRS N07223) and Commission Nationale de l’Informatique et des Libertés (CNIL N1245228) and was considered compliant with the General Data Protection Regulation and was approved by the institutional review committee of Institut Curie on April 2020. HEBON was approved by the Protocol Toetsingscommissie van het Nederlands Kanker Instituut/Antoni van Leeuwenhoek Ziekenhuis (PTC07.1611/2007NKI-2007-375). MUV was approved by the Ethikkommission der Medizinischen Universität Wien (2190/2019). IHCC was approved by the Komisja Bioetyczna Pomorskiego Uniwersytetu Medycznego w Szczecinie (Bioethics Committee of Pomeranian Medical University in Szczecin; BN-001/33/04).

