## Supplementary material for "Validation of the BOADICEA Model in a Prospective Cohort of *BRCA1/2* Pathogenic Variant Carriers"

### Methods

#### Censoring at risk-reducing salpingo-oophorectomy (RRSO)

BOADICEA does not consider the potential effect of risk-reducing salpingo-oophorectomy (RRSO) on breast cancer risk. To assess the possible impact on the results we considered RRSO as a censoring event in the analysis. This reduced the number of incident breast cancers by 48% (Table s1) and model performance estimates were associated with wide confidence intervals. Although there was an increase in the estimated AUC, there were larger deviations between the observed and expected numbers of cases in the individual quintiles of predicted risk compared to the analysis that ignored RRSO (Figure s3). The results suggest that RRSO should not be used as a censoring event when applying BOADICEA in *BRCA1/2* carriers in line with the lack of a pronounced effect of RRSO on breast cancer risk in published studies [1, 2].

Table s1 A summary of genetic and epidemiological characteristics of the eligible participants at baseline. Percentage was shown in women with information available.

| | Healthy women | Incident BC cases[a] | Incident DCIS cases[b] |
|---|---|---|---|
| **Number of participants, N** | | | |
| **Cohort-1** | 2770 | 186 | 23 |
| ***BRCA1* PV carriers** | 1487 | 116 | 11 |
| ***BRCA2* PV carriers** | 1283 | 70 | 12 |
| **Cohort-2** | 1613 | 171 | 20 |
| ***BRCA1* PV carriers** | 898 | 107 | 11 |
| ***BRCA2* PV carriers** | 715 | 64 | 9 |
| **PRS, mean (sd)** | | | |
| | 0.03 (1.04) | 0.31 (1.09) | 0.47 (0.73) |
| **Age at baseline, N (%)** | | | |
| **<30** | 492 (17.8%) | 17 (9.1%) | 1 (4.3%) |
| **[30,40)** | 847 (30.6%) | 53(28.5%) | 8 (34.8%) |
| **[40,50)** | 710 (25.6%) | 61 (32.8%) | 8 (34.8%) |
| **[50,60)** | 418 (15.1%) | 37(19.9%) | 5 (21.7%) |
| **[60,70)** | 243 (8.8%) | 17 (9.1%) | 1 (4.3%) |
| **≥70** | 60 (2.2%) | 1 (0.5%) | 0 (0.0%) |
| **median (IQR), years** | 42 (32-50) | 44 (36-52) | 42 (36, 50) |
| **Follow-up time, years** | | | |
| **mean (sd)** | 3.6 (1.4) | 2.7 (1.5) | 2.2 (1.3) |
| **Median (IQR)** | 4.0 (3.0-5.0) | 2.0 (1.0-4.0) | 2.0 (1.0-3.0) |
| **Age at menarche, N (%)** | | | |
| **<11** | 89 (3.5%) | 9 (5.6%) | 0 (0.0%) |
| **[11,12)** | 353 (14.0%) | 20 (12.5%) | 2 (10.0%) |
| **[12,13)** | 562 (22.4%) | 38 (23.8%) | 3 (15.0%) |
| **[13,14)** | 612 (24.3%) | 37 (23.1%) | 7 (35.0%) |
| **[14,15)** | 497 (19.8%) | 34 (21.2%) | 2 (10.0%) |
| **[15,16)** | 233 (9.3%) | 16 (10.0%) | 5 (25.0%) |
| **≥16** | 168 (6.7%) | 6 (3.8%) | 1 (5.0%) |
| **Missing** | 256 | 26 | 3 |
| **Menopausal status, N (%)** | | | |
| **Pre-menopausal** | 1715 (61.9%) | 118 (63.4%) | 14 (60.9%) |
| **Post-menopausal** | 1055 (38.1%) | 68 (36.6%) | 9 (39.1%) |
| **Age at menopause (among post-menopausal women), N (%)** | | | |
| **<40** | 230 (22.9%) | 9 (13.4%) | 3 (33.3%) |
| **[40,45)** | 231 (23.0%) | 18 (26.9%) | 2 (22.2%) |
| **[45,50)** | 248 (24.6%) | 19 (28.4%) | 3 (33.3%) |
| **[50,55)** | 257 (25.5%) | 18 (26.9%) | 0 (0.0%) |
| **≥55** | 40 (4.0%) | 3 (4.5%) | 1 (11.1%) |
| **Missing** | 49 | 1 | 0 |
| **Use of hormonal replacement treatment (among post-menopausal women), N (%)** | | | |

| | | | |
|---|---|---|---|
| **Current estrogen only type** | 97 (10.1%) | 7 (11.3%) | 0 |
| **Current other type** | 61 (6.4%) | 4 (6.5%) | 0 |
| **Former** | 169 (17.6%) | 4 (6.5%) | 1 (14.3%) |
| **Never** | 631 (65.9%) | 47 (75.8%) | 6 (85.7%) |
| **Missing** | 97 | 6 | 2 |
| **Parity, N (%)** | | | |
| **0** | 896 (32.4%) | 51 (27.4%) | 4 (17.4%) |
| **1** | 471 (17.1%) | 33 (17.7%) | 4 (17.4%) |
| **2** | 899 (32.6%) | 62 (33.3%) | 7 (30.4%) |
| **≥3** | 495 (17.9%) | 40 (21.5%) | 8 (34.8%) |
| **Missing** | 9 | 0 | 0 |
| **Age at first live birth (among parous women), N (%)** | | | |
| **<20** | 161 (8.7%) | 11 (8.3%) | 0 (0.0%) |
| **[20,25)** | 553 (29.9%) | 42 (31.6%) | 8 (42.1%) |
| **[25,30)** | 717 (38.7%) | 56 (42.1%) | 5 (26.3%) |
| **≥30** | 421 (22.7%) | 24 (18.0%) | 6 (31.6%) |
| **Missing** | 22 | 2 | 0 |
| **Use of oral contraceptive, N (%)** | | | |
| **Current** | 675 (25.7%) | 37 (21.6%) | 2 (10.0%) |
| **Former** | 1632 (62.2%) | 116 (67.8%) | 17 (85.0%) |
| **Never** | 317 (12.1%) | 18 (10.5%) | 1 (5.0%) |
| **Missing** | 146 | 15 | 3 |
| **Body Mass Index (kg/m2), N (%)** | | | |
| **<18.5** | 95 (3.5%) | 5 (2.7%) | 1 (4.5%) |
| **[18.5,25)** | 1561 (57.4%) | 109 (59.6%) | 12 (54.5%) |
| **[25,30)** | 679 (25.0%) | 50 (27.3%) | 4 (18.2%) |
| **≥30** | 382 (14.1%) | 19 (10.4%) | 5 (22.7%) |
| **Missing** | 53 | 3 | 1 |
| **Height (cm), N (%)** | | | |
| **<152.91** | 112 (4.1%) | 5 (2.7%) | 0 (0.0%) |
| **[152.91, 159.65)** | 372 (13.6%) | 25 (13.7%) | 6 (27.3%) |
| **[159.65, 165.96)** | 914 (33.4%) | 52 (28.4%) | 4 (18.2%) |
| **[165.96, 172.70)** | 824 (30.2%) | 66 (36.1%) | 7 (31.8%) |
| **≥172.70** | 511 (18.7%) | 35 (19.1%) | 5 (22.7%) |
| **Missing** | 37 | 3 | 1 |
| **Alcohol consumption (g/day), N (%)** | | | |
| **<5** | 1111 (43.1%) | 66 (37.5%) | 7 (36.8%) |
| **[5,15)** | 1003 (39.0%) | 75 (42.6%) | 6 (31.6%) |
| **[15,25)** | 272 (10.6%) | 19 (10.8%) | 5 (26.3%) |
| **[25,35)** | 122 (4.7%) | 9 (5.1%) | 0 (0.0%) |
| **[35,45)** | 45 (1.7%) | 5 (2.8%) | 1 (5.3%) |
| **≥45** | 22 (0.9%) | 2 (1.1%) | 0 (0.0%) |
| **Missing** | 195 | 10 | 4 |
| **Risk-reducing salpingo-oophorectomy, N (%)** | | | |
| **Cohort-1:** | | | |

| | | | |
|---|---|---|---|
| **Women with RRSO before the baseline** | 1070 (38.6%) | 83 (44.6%) | 12 (92.3%) |
| **Censored after the baseline** | 169 (6.1%) | 9 (4.8%) | 1 (7.7%) |
| **Cohort-2:** | | | |
| **Women with RRSO before the baseline** | 666 (41.3%) | 74 (43.3%) | 10 (90.9%) |
| **Censored after the baseline** | 116 (7.2%) | 8 (4.7%) | 1 (9.1%) |

[a]Incident breast cancer cases during the five-year prediction period.

[b]Incident ductal carcinoma in situ cases during the five-year prediction period.

PV: pathogenic variant; FH: family history.

Table s2: Calibration and discrimination of five-year predicted breast cancer risks using the cohort-2 samples (N=1,804) under the model considering pathogenic variant status in *BRCA1* and *BRCA2,* questionnaire-based risk factors, polygenic risk score and family history (FH). Model performance was examined by including information on all available relatives, or only first or second degree relatives.

| **Degrees of relatives included in the pedigree-based FH** | **Category** | **AUC** | **Harrell's C-index** | **E/O** | **Calibration slope** |
|---|---|---|---|---|---|
| 1st degree relatives only | All women | 0.79 (0.76, 0.82) | 0.70 (0.66, 0.74) | 1.01 (0.87, 1.17) | 1.03 (0.97, 1.09) |
| | *BRCA1* carriers | 0.78 (0.74, 0.82) | 0.68 (0.63, 0.74) | 1.01 (0.83, 1.22) | 1.02 (0.95, 1.10) |
| | *BRCA2* carriers | 0.79 (0.75, 0.84) | 0.72 (0.66, 0.79) | 1.01 (0.79, 1.30) | 1.03 (0.94, 1.12) |
| 1st and 2nd relatives only | All women | 0.79 (0.76, 0.82) | 0.70 (0.66, 0.74) | 1.05 (0.90, 1.22) | 1.05 (0.99, 1.11) |
| | *BRCA1* carriers | 0.78 (0.74, 0.82) | 0.69 (0.64, 0.73) | 1.04 (0.86, 1.25) | 1.04 (0.96, 1.12) |
| | *BRCA2* carriers | 0.79 (0.74, 0.84) | 0.71 (0.65, 0.78) | 1.07 (0.84, 1.37) | 1.06 (0.97, 1.15) |
| Full collected pedigrees | All women | 0.79 (0.76, 0.82) | 0.70 (0.67, 0.74) | 1.07 (0.92, 1.24) | 1.06 (1.00, 1.12) |
| | *BRCA1* carriers | 0.78 (0.74, 0.82) | 0.69 (0.62, 0.74) | 1.05 (0.87, 1.27) | 1.05 (0.97, 1.13) |
| | *BRCA2* carriers | 0.79 (0.75, 0.84) | 0.72 (0.66, 0.77) | 1.10 (0.86, 1.40) | 1.07 (0.98, 1.16) |

Table s3: Calibration and discrimination of five-year predicted breast cancer risks under the BOADICEA model using different risk factor combinations by censoring DCIS (ductal carcinoma in situ) as unaffected.

| **Model** | **Category** | **N.unaffected** | **N.BCs** | **AUC** | **Harrell's C-index** | **E/O** | **Calibration slope** |
|---|---|---|---|---|---|---|---|
| **using cohort-1** | | | | | | | |
| PV+QRFs+PRS | All women | 2793 | 186 | 0.78 (0.75, 0.81) | 0.69 (0.66, 0.73) | 0.94 (0.81, 1.10) | 0.99 (0.93, 1.04) |
| | *BRCA1* carriers | 1498 | 116 | 0.76 (0.71, 0.80) | 0.66 (0.61, 0.70) | 0.96 (0.79, 1.16) | 0.99 (0.91, 1.06) |
| | *BRCA2* carriers | 1295 | 70 | 0.80 (0.76, 0.85) | 0.74 (0.69, 0.80) | 0.91 (0.71, 1.17) | 0.99 (0.91, 1.08) |
| **using cohort-2** | | | | | | | |
| PV+QRFs+PRS+FH | All women | 1633 | 171 | 0.79 (0.76, 0.82) | 0.70 (0.66, 0.74) | 1.18 (1.01, 1.38) | 1.11 (1.05, 1.17) |
| | *BRCA1* carriers | 909 | 107 | 0.78 (0.73, 0.82) | 0.68 (0.62, 0.73) | 1.15 (0.94, 1.40) | 1.09 (1.01, 1.17) |
| | *BRCA2* carriers | 724 | 64 | 0.80 (0.75, 0.85) | 0.73 (0.68, 0.77) | 1.24 (0.95, 1.61) | 1.13 (1.03, 1.23) |

PV: pathogenic variant status in *BRCA1* and *BRCA2*; QRFs: questionnaire-based risk factors; PRS: polygenic risk score; FH: family history

Table s4: Calibration and discrimination of five-year predicted breast cancer risks using the cohort-2 samples (N=1,804) under the full model considering pathogenic variant status in *BRCA1* and *BRCA2,* questionnaire-based risk factors, polygenic risk score and family history by age group.

| Age | N.Unaffected | N.BCs | AUC | Harrell's C-index | E/O | Calibration slope |
|---|---|---|---|---|---|---|
| < 50 years | 1190 | 139 | 0.80 (0.77, 0.84) | 0.72 (0.66, 0.75) | 1.00 (0.84, 1.19) | 1.03 (0.96, 1.09) |
| ≥ 50 years | 423 | 52 | 0.75 (0.67, 0.82) | 0.64 (0.55, 0.71) | 1.28 (0.96, 1.71) | 1.16 (1.03, 1.29) |

Figure s1: Consort diagram summarising the TRANsIBCCS cohort data

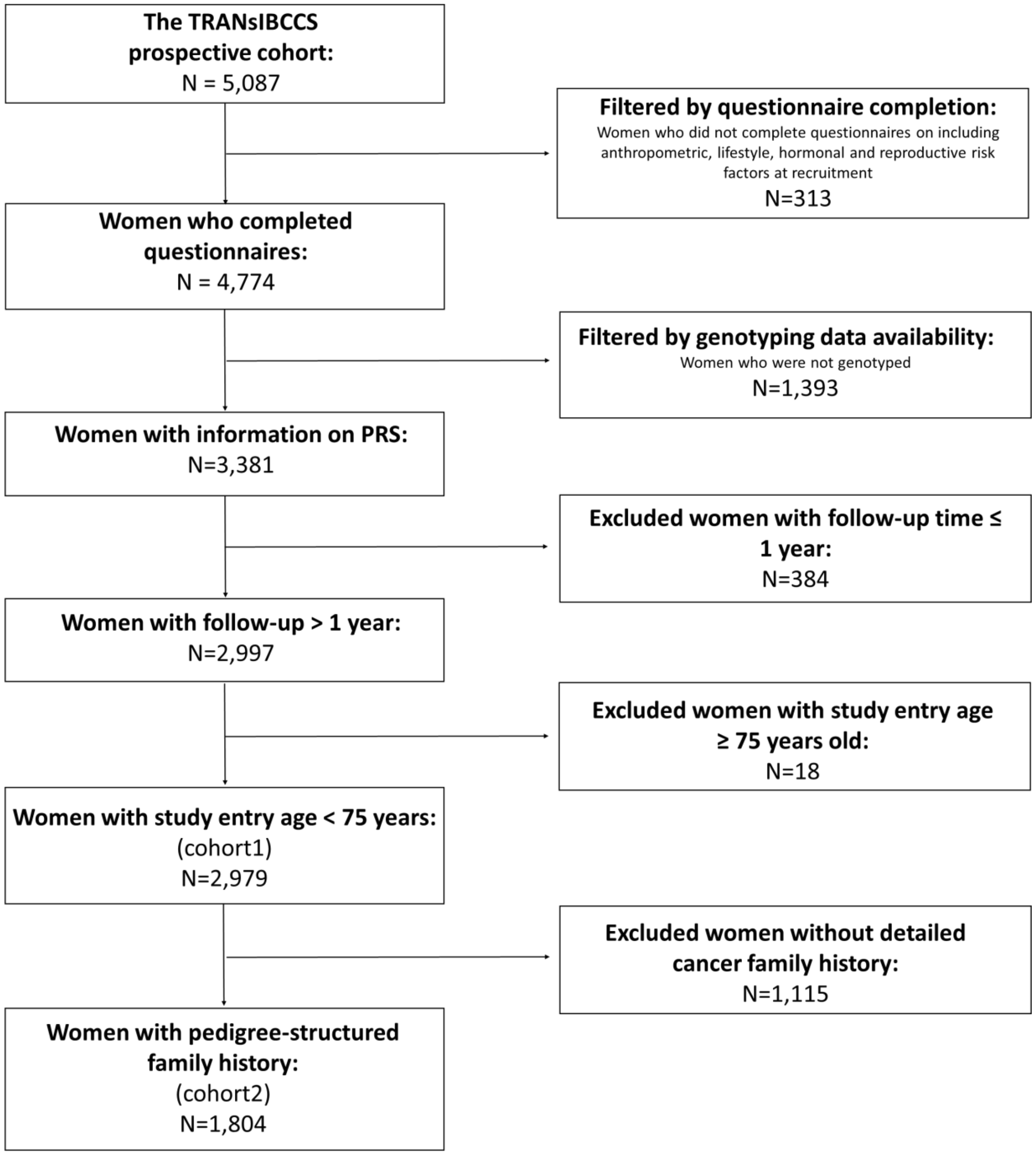

Figure s2: Observed and expected five-year breast cancer risks in quintiles of predicted risks, using the cohort-2 samples (N=1,804) under the model considering pathogenic variant status in *BRCA1* and *BRCA2,* questionnaire-based risk factors, polygenic risk score and family history. Model performance was examined by considering (a) only 1$^{st}$ degree relatives, (b) 1$^{st}$ and 2$^{nd}$ degree relatives and (c) the full collected pedigrees including more distant relatives. The dashed line is the diagonal line with slope equal to 1 (corresponding to E/O ratio of 1 for each quintile).

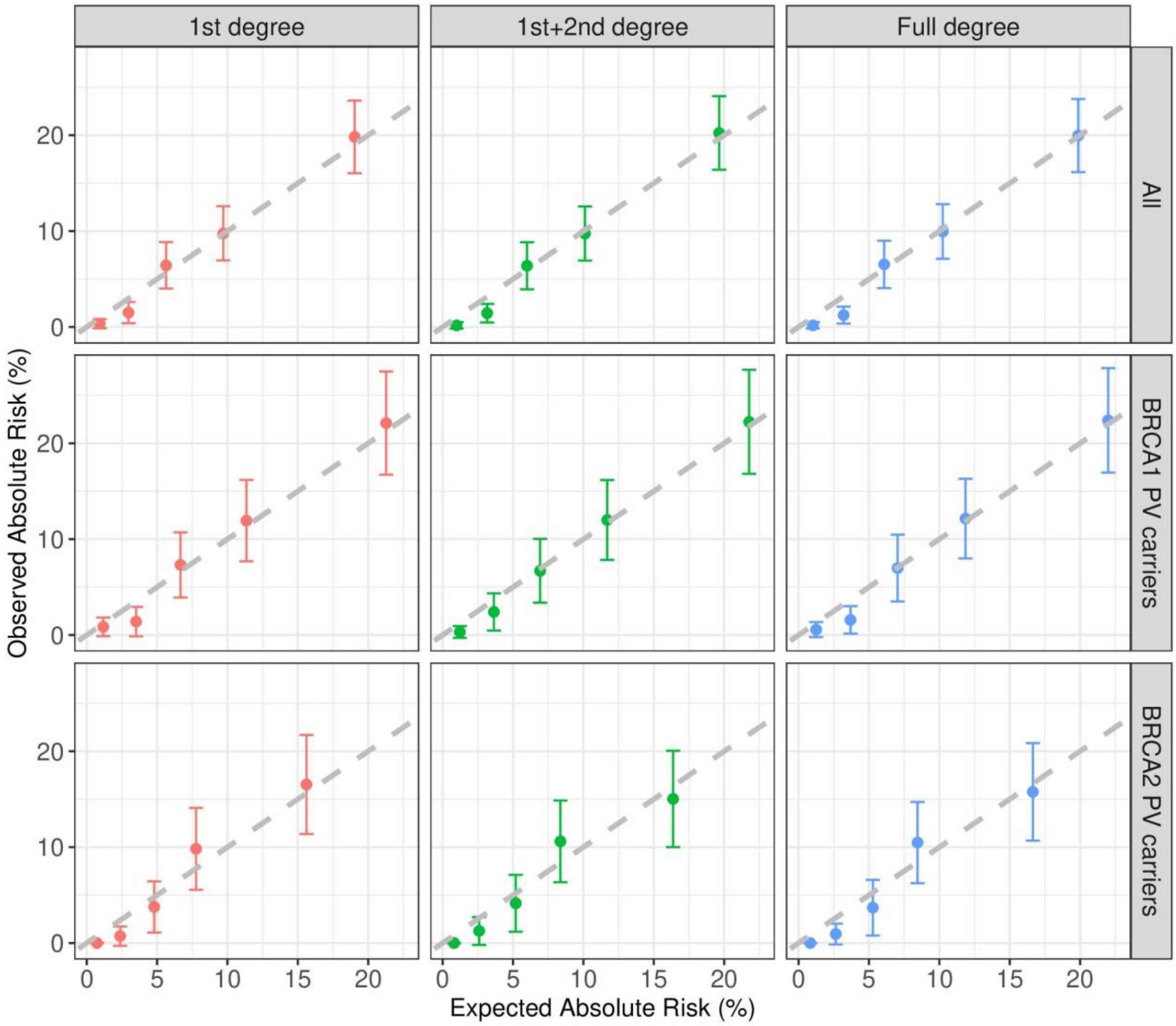

Figure s3: Observed and expected five-year breast cancer risks in quintiles of predicted risks, using (1) the cohort-1 samples (N=2,979) under the model considering PV, QRFs and PRS; (2) the cohort-2 samples with FH information (N=1,804) under the model considering PV, QRFs, PRS and FH by censoring DCIS (ductal carcinoma in situ) as unaffected. The dashed line is the diagonal line with slope equal to 1 (corresponding to E/O ratio of 1 for each quintile). PV: pathogenic variant status in *BRCA1* and *BRCA2*; QRFs: questionnaire-based risk factors; PRS: polygenic risk score; FH: family history.

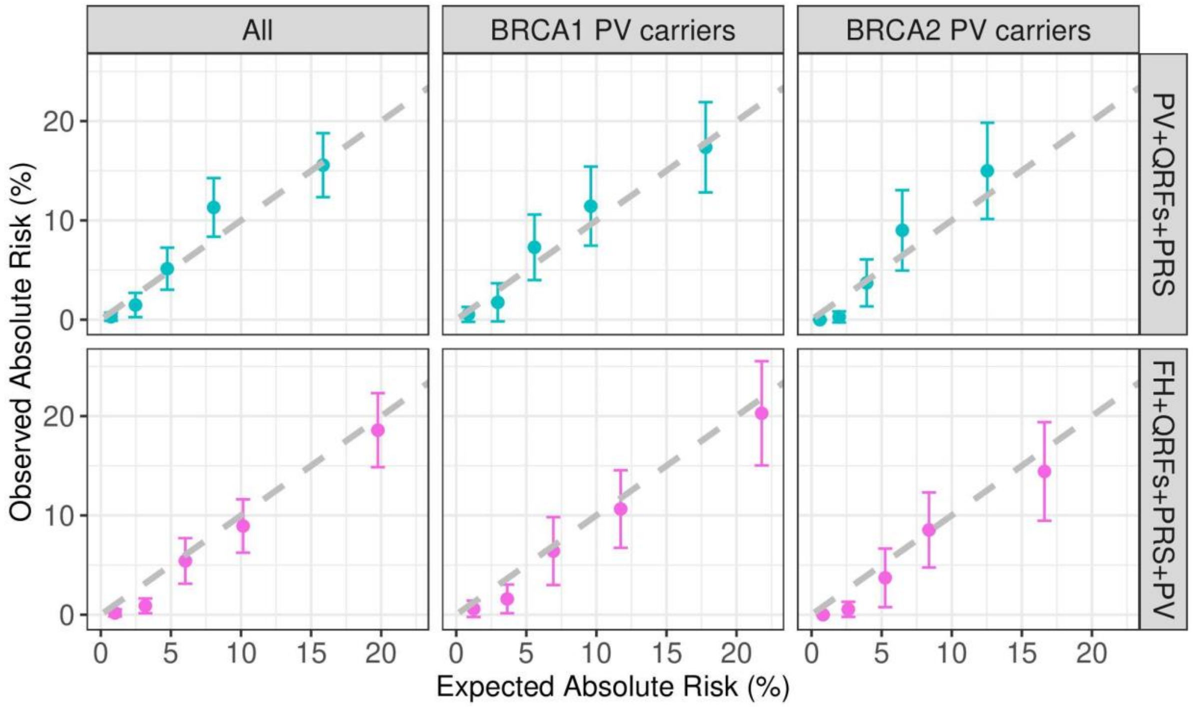

Figure s4: Observed and expected five-year breast cancer risks in quintiles of predicted risks, using the cohort-2 samples when censoring at risk-reducing salpingo-oophorectomy (N=1,054 eligible at baseline) under the model considering PV, QRFs, PRS and FH. The dashed line is the diagonal line with slope equal to 1 (corresponding to E/O ratio of 1 for each quintile). PV: pathogenic variant status in *BRCA1* and *BRCA2*; QRFs: questionnaire-based risk factors; PRS: polygenic risk score; FH: family history.

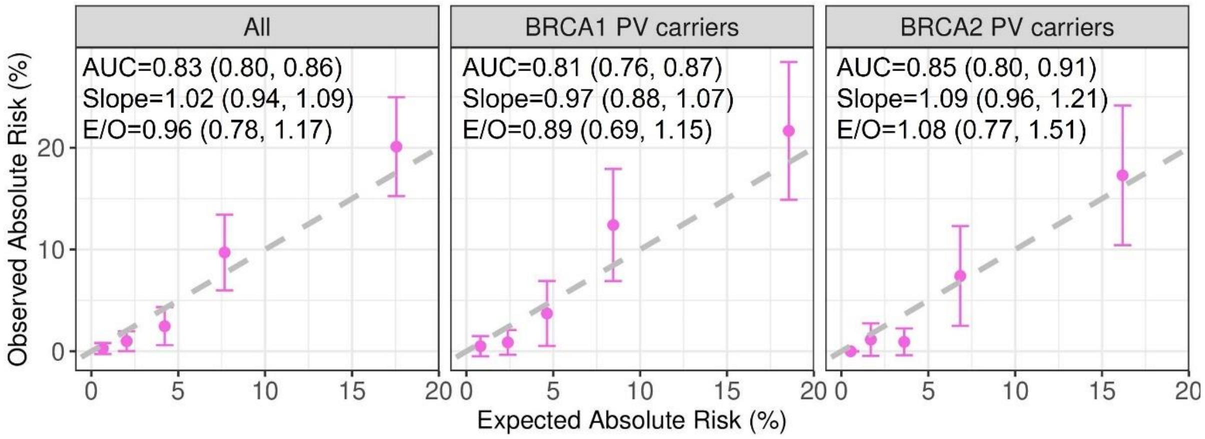

Figure s5: Observed and expected five-year breast cancer risks in quintiles of predicted risks, using the cohort-2 samples (N=1,804) under the model considering pathogenic variant status in *BRCA1* and *BRCA2*, questionnaire-based risk factors, polygenic risk score and family history by age group.

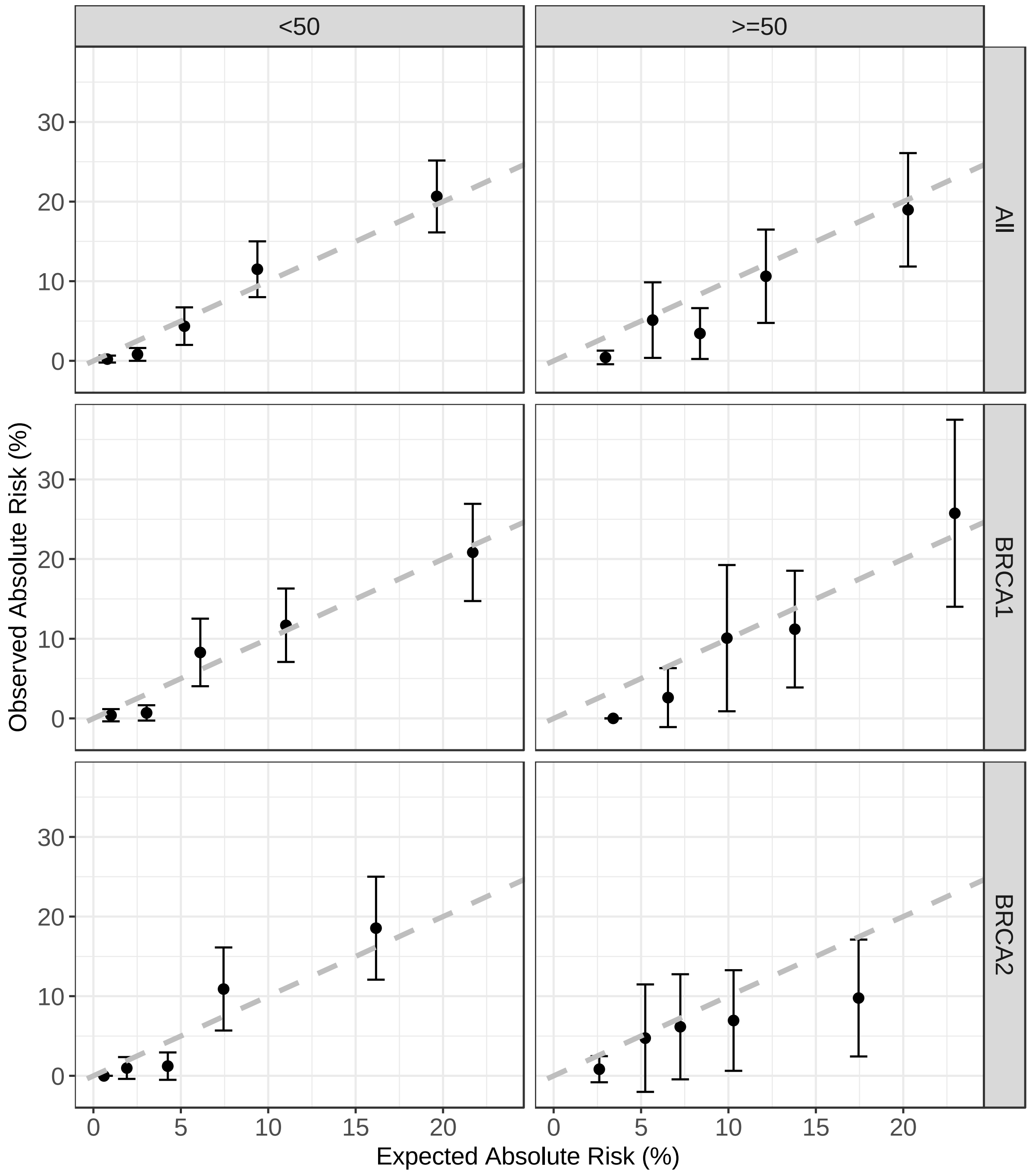
